# Seasonal Disease in the United States Has the Hallmarks of an Entrained Circannual Clock

**DOI:** 10.1101/2021.05.26.21257655

**Authors:** Andrew F. Schober, Andrey Rzhetsky, Michael J. Rust

## Abstract

Infectious human diseases are often characterized by seasonal variation in incidence which produces recurring annual peaks with precise timing. Changing weather conditions such as temperature and humidity are known to affect virus transmission, but we consider the possibility that an intrinsic rhythm in susceptibility, entrained to annual cues, also contributes to seasonality. These two mechanisms produce different predictions about how the timing and amplitude of seasonal disease depend on environmental forcing. Using databases of health insurance claims and weather across U.S. counties, we find that the timing of winter-peaking diseases, but not their amplitude, scales with latitude. This latitude scaling is shared across many diagnosis codes involving multiple pathogens. Regression against simple models suggests an underlying limit cycle that, in temperate zones, is entrained to annual changes in irradiance, possibly a human circannual clock.

**Significance statement:** Peak timing but not amplitude of seasonal disease scales with latitude, suggesting a circannual rhythm in host susceptibility.

## Main Text

Seasonal variation in incidence is a common feature of infectious human disease (1-5) which has been documented even prior to the discovery of microbes (6, 7). However, the fundamental mechanisms underlying seasonal disease remain incompletely understood. Much work has sought to identify direct drivers of seasonal disease, whereby changing external weather conditions alter either pathogen transmission (8-10), survival (11-17), host behavior (18-21), or immune response (22). Decreased temperature, humidity, and exposure to sunlight have been shown statistically to precede the onset of seasonal respiratory infection in temperate zones (23-28). However, attempts to demonstrate the effect of such conditions on the susceptibility of human subjects to rhinovirus have been inconclusive (29-31). Furthermore, it is difficult to reconcile these explanations of seasonality with the significant burden of respiratory illness in the tropics, where some seasonal variation is observed despite the warm and humid climate conditions year-round (32-37).

The search for direct environmental drivers of disease risk implicitly assumes that seasonality results from the external forcing of a system that would otherwise rest at a steady state. Here we consider an alternative hypothesis: that the underlying biology may be intrinsically oscillatory, so that disease susceptibility depends in part on the internal state of the host. Self-sustaining oscillations are widespread throughout biology, including circadian clocks (38-40), the menstrual cycle (41), circatidal rhythms (42, 43), and circannual rhythms (44-52). Though most disease models have not considered internal oscillator mechanisms, it has been previously proposed that the seasonal pattern of infectious disease might reflect an endogenous physiological cycle in humans (32, 53, 54). These two classes of mechanisms — external forcing versus the entrainment of a self-sustaining oscillator — produce qualitatively different predictions about how the timing or amplitude of seasonal disease will respond to the relative strength of environmental inputs. While it is not feasible to do controlled laboratory experiments on annual rhythms in humans, the existence of large-scale health insurance claims data sets (55) coupled with detailed weather records from across the U.S. (56) allow us to quantitively study how these features of seasonal disease scale with climate and geography.

Driven systems, analogous to an adult pushing a child on a swing, have a stable steady state in the absence of external forcing. When forcing is weak, one can approximate a driven oscillator with linearized dynamics (see Supplementary text). Two qualitative features of the driven response emerge. First, the amplitude of the response is proportional to the input strength (Fig. 1A). Second, the phase of the output oscillation is not dependent on input strength. In contrast, self-sustaining oscillators are described by limit cycles. In the swingset analogy, the child pumps the swing herself and reaches a defined amplitude without external pushes. The simplest model of a weakly driven limit cycle is a phase oscillator where external input modulates the angular velocity (57). This model can be solved analytically with the following general results: if the natural period of the oscillator and the driving period are mismatched, stable entrainment only occurs if the amplitude of the input is sufficiently large (see Supplementary text). The phase of an entrained oscillator varies inversely with input strength as the entrainment boundary is approached (Fig. 1A). Thus, a self-sustaining oscillator mechanism implies a nearly constant amplitude, a phase that depends on input strength, and the presence of an entrainment boundary. To discriminate between these possibilities — the response of a stable system to oscillatory forcing vs. an underlying limit cycle oscillator— we studied how seasonal disease timing scales with the properties of the environment across the United States.

**Figure 1.**
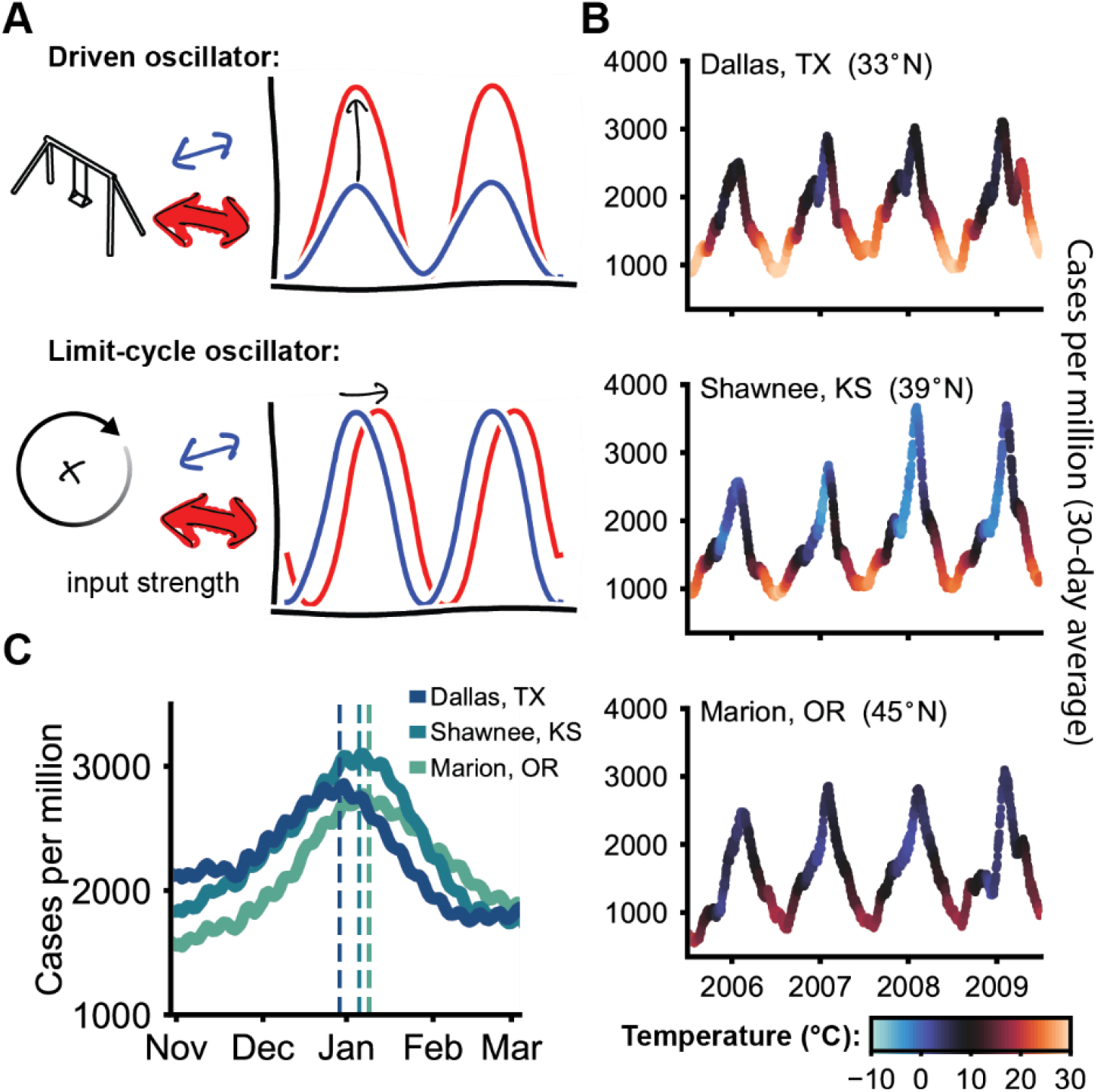
How seasonal disease scales with increasing input strength can be used to distinguish the underlying mechanism. **(A)** Two qualitatively different mechanisms can convert an oscillatory input (the environment) to an oscillatory output (seasonal disease). Driven systems have a steady state in a constant environment but can be driven to oscillate by external forcing (like a swing-set). In contrast, limit cycles produce spontaneous oscillations with fixed amplitude but can entrain to inputs that are close in frequency; a common example is the circadian clock. When these systems are weakly driven, varying input strength changes the amplitude of the driven oscillator without affecting its phase. Conversely, increased input strength shifts the phase of a limit cycle oscillator without affecting its amplitude. **(B)** Three representative U.S. counties show similar amplitudes of winter-peaking seasonal disease despite geographic and climate differences. Case counts and temperature values reflect 30-day averages in each county. **(C)** Peak day is estimated from 4 years (April 2005 – April 2009) of 30-day averaged incidence rates, indicated on the plot by a dashed line. The pictured counties anecdotally illustrate a trend in which the peak day occurs later at higher latitudes.

## Results

We analyzed health insurance claims and climate conditions across 1,707 diverse U.S. counties. Our analysis combines 73 of the most common winter-peaking diagnoses (Table S1), which are infections of the upper and lower respiratory tracts and related conditions. Disease timing and amplitude were computed from 30-day averaged incidence rates across a sample of 4 winter seasons, comprising a total of more than 68 million diagnoses. Representative plots of disease incidence and temperature for three geographically disparate U.S. counties are displayed in Fig. 1B. Similar amplitudes of seasonal infection can be observed across the diverse geography and climates of the continental U.S., with peaks occurring at very different temperatures in different locations. We computed the mean value and standard errors of peak day and amplitude (as peak/trough ratio) across the 4-year sample (Fig. 1C). Notably, we observe low year-to-year variation in both quantities (Dataset S1). Standard errors associated with the peak days (dashed lines) in Fig. 1C are less than 3 days. Thus, the aggregate dynamics of seasonal disease on annual timescales reflect a highly deterministic underlying process rather than stochastic bursts of infection. Lastly, the pictured counties demonstrate a general trend found in peak timing across the country; peak day occurs systematically earlier at lower latitudes. Notably, the scaling of phase with latitude has been established as a general feature of circannual rhythms in animals, which are entrained to photoperiodic cues (44, 49-52).

Plotting average peak day by county onto a map of the United States reveals the relationship between geography and phase (Fig. 2A). The density map in Fig. 2B shows the approximately linear trend relating latitude to peak day. Consequently, the statistical association between peak day and latitude is highly significant (*R* = 0.44, *p* =2.18× 10^−81^, linear slope *m*= 0.83). To assess whether geographic or climate factors are most predictive of peak day, we devised a test using *L*_1_ regularized regression (LASSO). A series of LASSO fits predicting peak day from geographic, climatological, and demographic covariates were computed under various regularization strengths (Fig. 2C). As the *L*_1_ penalty increases, most coefficients tend toward zero, revealing which variables provide the greatest predictive power. The climate variables considered were the temperature, specific humidity, and solar irradiance of each county in the month preceding the annual peak, which have been implicated by prior studies as drivers of seasonal disease (23-25, 28, 36). However, the results of our LASSO fits indicate that latitude is the best predictor of peak timing. Latitude alone explains ∼20% of the observed variance (Fig. 2C, column 4), while the inclusion of climate factors only improves the model by an additional 10% (column 1). Mean peak day within geographic bands increases monotonically with latitude (Fig. 2D).

**Figure 2.**
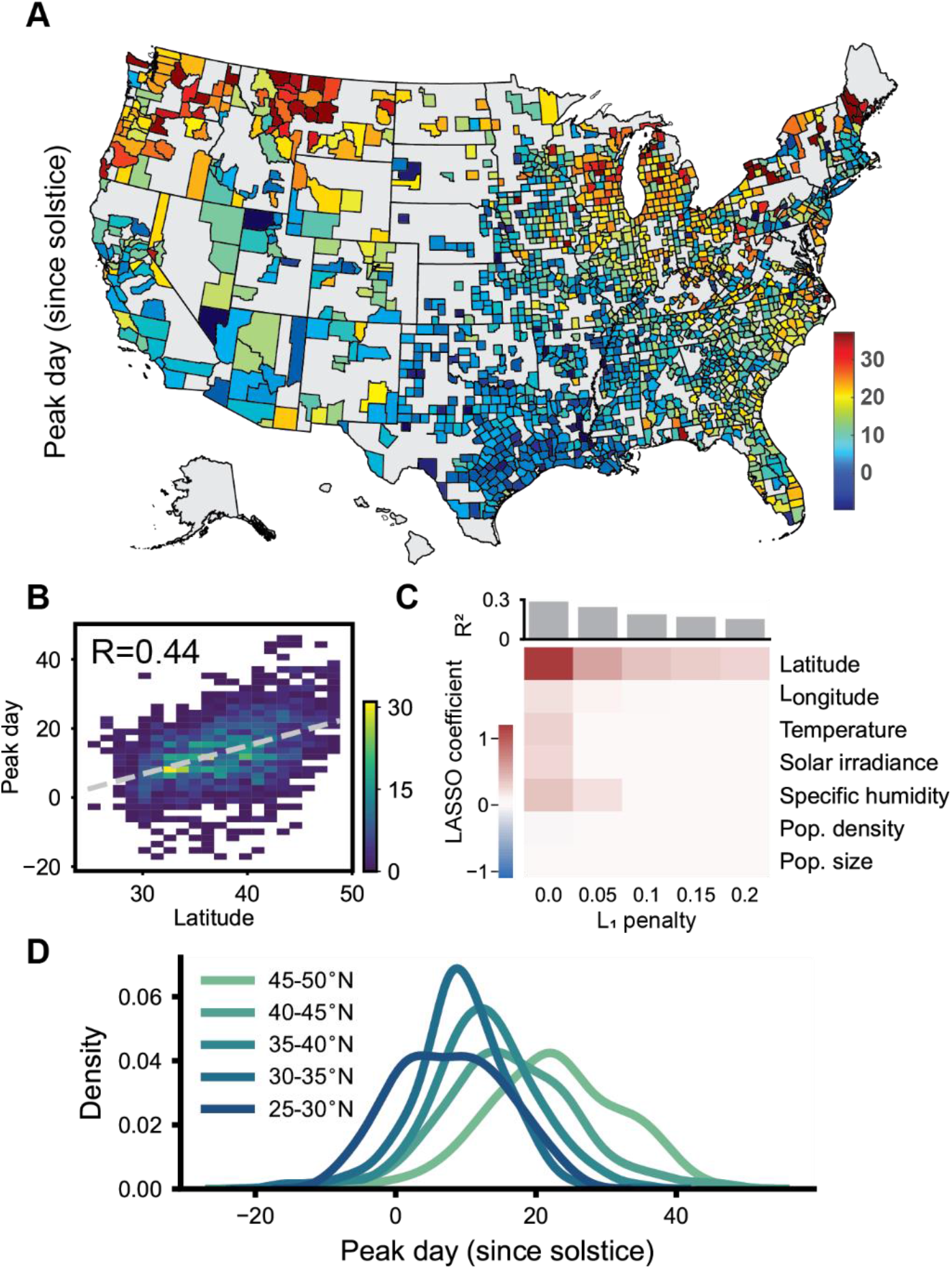
The timing of winter-peaking seasonal disease in the U.S. is most determined by latitude, not climate. **(A)** Average peak timing (days since winter solstice) of winter-seasonal disease shows a clear geographic trend. Peak day was computed individually for 1,707 U.S. counties across a 4-year sample and demonstrated precise timing. **(B)** Geographic latitude was found to be the strongest correlate with peak day. Pixel intensity indicates the number of counties. The linear fit line with a slope of 0.83 is shown in grey dashes; the Pearson correlation coefficient is displayed along the bottom right. **(C)** LASSO models of peak day indicate that latitude has more explanatory power than climate variables. Each column displays the coefficient of determination *R*^2^ (bar graph) and coefficient values (heatmap) for a series of LASSO fits computed at a different regularization strength or *L*_1_ penalty. Coefficient magnitudes indicate the relative predictive power of each variable at a given regularization. Increasing the *L*_1_ penalty represents an accuracy-simplicity tradeoff and favors latitude as all other coefficients tend to zero. **(D)** Mean peak day increases monotonically with distance from the equator across latitude-bands.

We sought to address whether this phase-latitude trend is due to a specific diagnosis or specific pathogen which might dominate seasonal trends. In fact, the linear slope of the phase-latitude trend is robust even when the top 10 most common diagnoses, comprising over 70% of the dataset, are removed (Figure S1A). We also detected a consistent phase-latitude slope when analyzing only diagnoses for streptococcal sore throat, the most common code associated with a specific pathogen (Figure S1B). Thus, we conclude that dependence of seasonal disease timing on latitude is unlikely to be caused by idiosyncratic properties of a specific pathogen but is instead a shared feature of many seasonal conditions.

Compared to the timing of annual peaks, disease amplitude does not vary strongly with latitude (Fig. 3A). This suggests that the prevalence of winter-peaking diseases is not directly linked to the severity of winter weather (e.g. cold, dry air) which varies markedly across the U.S. The statistical association between latitude and amplitude is comparatively weaker (Fig. 3B, *R* = 0.07, *p* = 3.07 × 10^−3^). Our LASSO models indicate that the weak association between latitude and amplitude may arise from collinearity with demographic factors such as population size (Fig. 3C). The larger amplitude in rural, less-populous counties may derive from behavioral, socioeconomic, or diagnostic differences. Taken together, the clear phase-latitude relationship and relative lack of amplitude scaling with geography are consistent with the hypothesis that an underlying human circannual clock affects disease susceptibility; these scaling behaviors are hallmarks of an entrained self-sustaining oscillator.

**Figure 3.**
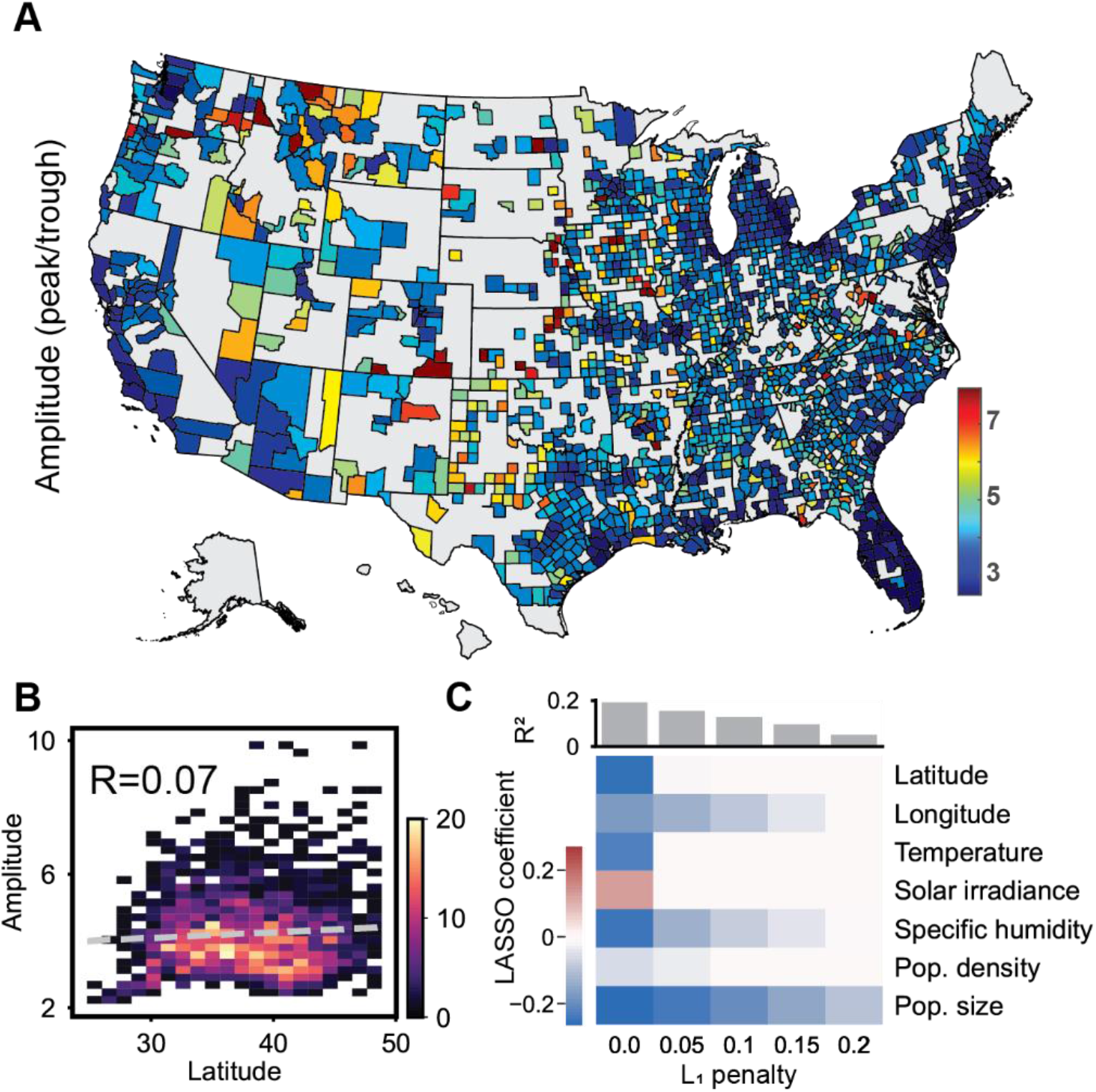
The amplitude of winter-peaking seasonal disease in the U.S. is only weakly influenced by climate and geography. **(A)** Average amplitude (peak/trough ratio) was computed individually for 1,707 U.S. counties across a 4-year sample and demonstrated low year-to-year variance. **(B)** Disease amplitude has a comparatively modest linear association with latitude. The linear fit line with a slope of 0.02 is shown in grey dashes; the Pearson correlation coefficient is displayed along the bottom right. **(C)** LASSO models of disease amplitude reveal that peak/trough ratio is enriched in arid, rural counties with low population size. Columns display the coefficient of determination *R*^2^ (bar graph) and coefficient values (heatmap) for a series of LASSO fits computed at a different regularization strength or *L*_1_ penalty. Coefficient magnitudes indicate the relative predictive power of each variable at a given regularization.

Can a limit cycle oscillator with environmental time-series as inputs recapitulate disease timing better than a driven harmonic oscillator? To test this, we regressed the disease incidence in each county against generic models of driven system and a self-sustaining oscillator (see Supplementary text). Model inputs were regressed to an optimal linear combination of temperature, specific humidity, and solar irradiance derived from time series weather data for each county. Using the models, we assessed which mechanism was best able to explain the observed phase-latitude trend. In the health insurance claims data set, peak day increases at a rate of 0.83 days per degree latitude (Fig. 4A). Peak days predicted by the best-fit driven model, which favored temperature and specific humidity as inputs (Table S2), only constitute an increase of 0.06 days per degree latitude (Fig. 4B, *R* = 0.09, *p* = 1.64 × 10^−4^). This finding suggests that the phase differences observed across latitudes cannot be explained solely by local weather time-series. In comparison, peak days predicted by the phase oscillator exhibit a dependence on latitude similar to the observed trend (Fig. 4C, *R* = 0.87, *p* < 1.80 × 10^−308^), and the best fit for its input function is only dependent on solar irradiance; coefficients for other climate variables tended toward zero (Table S2). This demonstrates that a limit cycle mechanism can recapitulate the observed timing using a single input proportional to photoperiod, an established entraining cue of known circannual clocks (49-52). Finally, the phase oscillator model can be extrapolated to predict the presence of an entrainment boundary near 18°N degrees, within the tropics.

**Figure 4.**
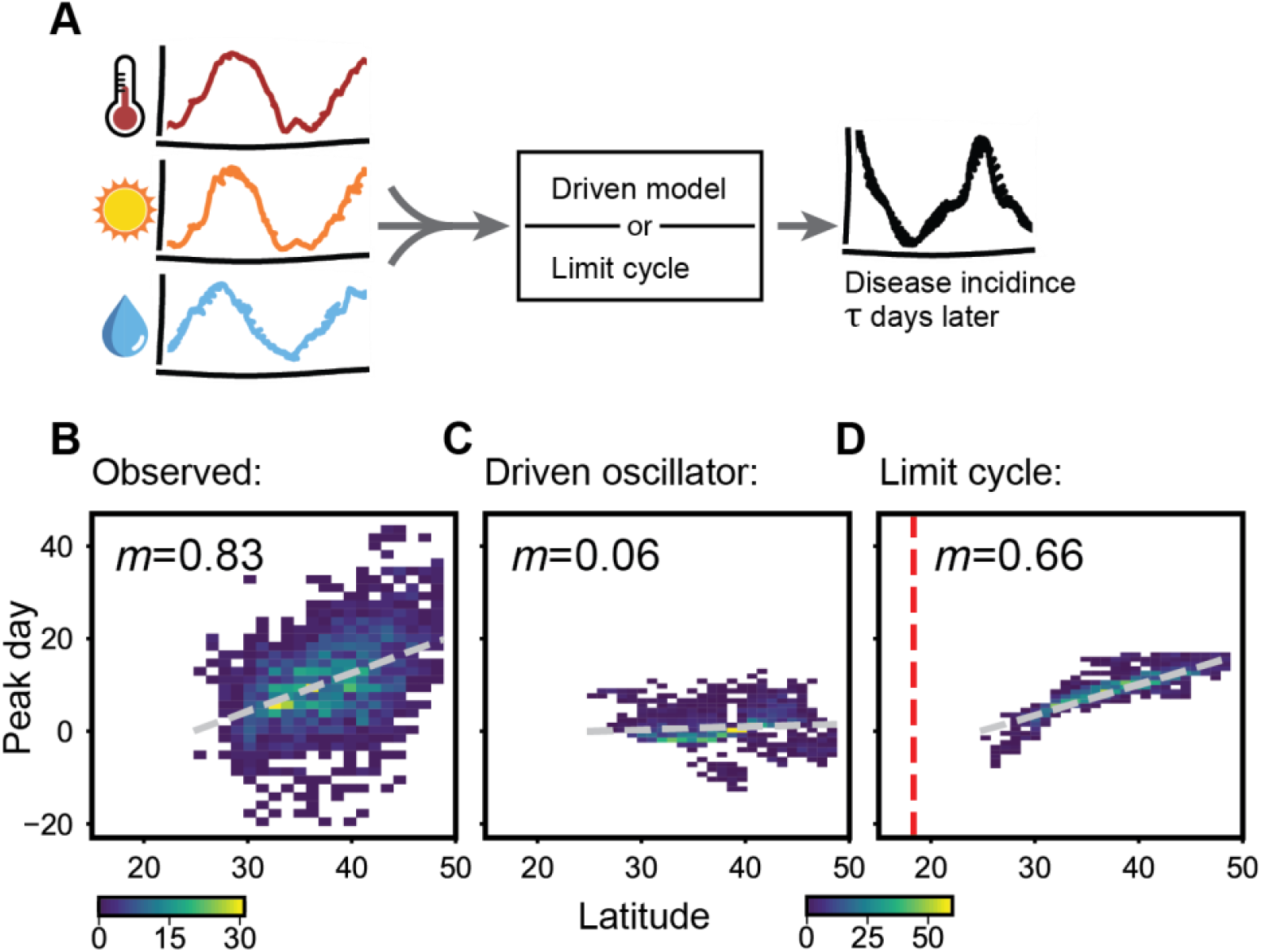
Simple models suggest that the timing of winter-seasonal disease can be described by an underlying limit cycle entrained to seasonal cues. **(A)** Schematic of regression analysis used to predict disease incidence from time-lagged weather data consisting of temperature, solar irradiance, and specific humidity. For each model, nonlinear least-squares optimization was used to find a single set of best fit parameters relating weather time-series to future disease incidence in 1,707 U.S. counties across a 4-year sample. **(B)** The observed peak day versus latitude relationship compared to those predicted by generic driven oscillator and limit cycle oscillator models. Linear fit line is shown in grey dashes and the slope is labeled along the top of each plot. Each plot is offset such that peak day 0 corresponds to the linear fit of the southernmost county. **(C)** Because the driven linear oscillator can only recapitulate phase differences that are already present in its environmental inputs, it is unable to fully explain the observed phase-latitude relationship. (**D)** The limit cycle oscillator can reproduce the relationship between latitude and peak day seen in the data. Dashed red line indicates the predicted limit of entrainment in the model (approximately 18°N).

## Discussion

The two mechanisms considered here, direct response of a system to environmental forcing and the entrainment of a self-sustaining oscillator, are not mutually exclusive. A complete picture of disease dynamics likely involves both classes of mechanisms. Nevertheless, the success of a solar irradiance-entrained limit cycle model in predicting winter respiratory disease timing motivates further inquiry into the possibility of a circannual rhythm in disease susceptibility.

In light of this hypothesis, we also revisit existing data on the seasonality of human infectious diseases. A prior analysis of influenza in Brazil found that annual peak timing in the sub-tropical region was delayed by upwards of one month as latitudinal distance from the equator increased, similar to the phase gradient we observe (33). Consistent with other studies (34, 37, 58, 59), the fluctuation of influenza incidence was comparatively weak and irregular in the tropical part of Brazil. The geographic boundary delineating these two disparate dynamics was identified as 16°S latitude, which is consistent with the entrainment boundary predicted by our model. More recently, this analysis has been extended to demonstrate a similar latitudinal gradient for influenza and respiratory syncytial virus spanning the temperate zones of both hemispheres (35). A study of influenza-like-illness in Ho Chi Minh City, Vietnam (11°N latitude) identified nonannual dynamics with a dominant periodicity of ∼200 days (37). Just outside of an entrainment boundary, the period of an oscillator is not constant and may slip between its natural period and the driving period; thus, the presence of nonannual fluctuations in tropical climates may also suggest an underlying circannual clock.

Latitudinal gradients in disease timing are not limited to winter-peaking diseases. For example, nonpolio enterovirus infections in the U.S. were found to follow a similar latitudinal gradient in peak timing distributed between July and August (60). Prior to its eradication, the timing of poliomyelitis cases mirrored the latitudinal gradient produced by nonpolio enteroviruses but delayed such that the mean timing in each state occurred about one month later. While there are many differences between summer enterovirus infections and winter respiratory diseases, their qualitatively similar gradients in timing might be explained in part by an intrinsic rhythm in host physiology, inducing peak susceptibility to different diseases at different points in the annual cycle, rather than in terms of the unique transmission properties and environmental drivers of each disease. However, more work is needed to understand the role of transmission in setting the timing of seasonal disease in both tropical and temperate climate contexts.

While we favor an interpretation that near-annual limit cycle oscillator with a human circannual clock, another possibility is a self-sustaining oscillator that emerges from the epidemiology of infection, recovery, and subsequent fading of immunity. It is known that epidemiological models are also able to generate oscillations through the inclusion of seasonal forcing via periodic transmission rates (61-64), demographic fluctuations (65, 66), or constraints on healthcare intervention (67). The natural frequency of such models can vary widely depending on the birth and death rate of the population as well as the basic reproductive number of the pathogen. While hypothetical epidemiological rhythms for different diseases with a variety of natural frequencies could all be entrained to an annual cycle with a sufficiently strong input, in general such a scenario would result in a diverse spread of phase-latitude slopes for different diseases, some with opposite sign.

The potential existence of a circannual rhythm in human disease susceptibility could provide a basis for understanding why many diverse pathogens peak annually in temperate climates. Not only could these insights be used to improve models of seasonal disease, but they should also inspire further experimental investigation into physiological evidence for a circannual clock. Seasonal fluctuations in lymphatic organ size and immune function are well-established (68, 69). Recently, a human endocrine circuit was described that could explain seasonal changes in gland-size and the production of key hormones such as prolactin (70). The long-term regulation of prolactin is notably a feature of the sheep circannual clock (49). There may be other aspects of human physiology and social behavior that are linked to circannual rhythms in currently underappreciated ways; a better understanding of an underlying annual cycle in humans would have far-reaching impacts for basic biology, public health, and infectious disease epidemiology.

## Materials and Methods

### Data and code availability

Python code and aggregated diagnosis/weather data can be downloaded at github.com/afschober/seasonaldisease. Summary statistics of peak day and amplitude are provided as a part of the Supplementary Information.

### Description of data sets used

Health insurance claims and enrollees data for the years 2005 through 2009 were obtained from the Truven MarketScan database (55). Corresponding weather time-series were downloaded from Physical Solar Model (PSM) v3 hosted on The National Solar Radiation Database (NSRDB) website (56). Specific humidity was estimated from relative humidity by using the August-Roche-Magnus formula to calculate saturation vapor pressure from the temperature record. Pressure and relative humidity records from the NSRDB were then used to get the mass mixing ratio of water vapor in air. Population sizes for each county were obtained from the U.S. Census Bureau 2010 Census (71). Statistical analyses and nonlinear regressions were conducted using a custom Python script supported by the open-source SciPy (72) and statsmodels (73) modules.

### Identification of winter-peaking diseases

Winter-peaking diseases were selected through an initial analysis of peak day and amplitude applied to nationwide counts of each diagnosis in the database. Claims data were first normalized by the total number of enrollees in each year then smoothed using a 30-day average. Diagnoses with peaks near the winter-solstice were included if they satisfied the following empirically chosen criteria: Nationwide estimates of amplitude (peak/trough ratio) were required to exceed a 1.5 fold-increase with a standard error of less than 0.5. Standard errors of the corresponding peak times were also required to be less than 15 days. Health insurance claims that met the criteria but were not a sign of infectious disease, such as routine vaccinations, were removed manually to obtain a final list of diagnosis codes for study (Table S1).

### County estimates of peak day and amplitude

County-specific time-series of seasonal disease were constructed by normalizing the total claims counts of the selected diagnoses (Table S1) by the corresponding number of enrollees for each year of data considered. Time-series of incidence rate were then smoothed using a 30-day backward average. Peak day and amplitude as peak/trough ratio in each county were estimated across a sample of 4 winter seasons spanning from April 2005 to April 2009. Because photoperiod is an established entraining cue of known circannual clocks (44, 49-52), the winter solstice was adopted as a reference point for peak day. Thus, peak timing was expressed in units of days since the winter solstice. Amplitude was estimated as the average ratio between the annual peak incidence rate and the preceding summer’s trough. Peak day and amplitude were first estimated in all 3,122 U.S. counties for which both health insurance claims and weather data were available. To account for limited sampling depth, counties with an outlier (more than three standard deviations from the mean) in peak day or amplitude were removed from the dataset. The collection of counties considered was further trimmed based on the following empirically determined noise criteria: The standard error of peak day was required to be less than 15 days and the standard error of amplitude was required to be less than 1.5. The resulting dataset is comprised of 1,707 U.S. counties for which both peak day and amplitude could be precisely determined. Summary statistics for these counties are provided in Dataset S1. In our analysis of a subsample of less common diagnoses (Figure S1A), the same noise thresholds were applied to reach a total of 1,183 counties. For the purpose of analyzing the peak timing of streptococcal sore throat alone (Figure S1B), the noise thresholds were relaxed to 25 days of standard error in peak timing and a 5-fold standard error of amplitude, which resulted in the inclusion of 476 counties.

### LASSO models

*L*_1_ regularized regression (LASSO) models were built for countywide peak day and amplitude data against an assortment of geographic, climatological, and demographic factors across an array of regularization strengths. Temperature, solar irradiance, and specific humidity in each county were represented by their average value during the month of December, which encompasses the days leading up to the first peaks of winter-seasonal disease. Peak day, amplitude, and their covariates were first statistically standardized by centering their means and normalizing by their standard deviations. Then, LASSO coefficients and the corresponding goodness-of-fit statistics at each regularization strength were calculated using the statsmodels Python module (73).

### Regression of disease time-series against oscillator models

Disease time-series were regressed against simple models of a driven oscillator and a phase oscillator using nonlinear least squares. The details of each model are provided in the Supplementary text. Nonlinear regressions were computed using the trust region reflective algorithm provided by the SciPy Python module (72).

## Author contributions

A.F.S., A.R. and M.J.R. designed the study. A.F.S. performed the data analysis. A.F.S. and M.J.R. wrote the paper. All authors read and edited the paper.

## References

1. N. D. Noah, Cyclical patterns and predictability in infection. Epidemiol Infect 102, 175–190 (1989).

2. S. Altizer et al., Seasonality and the dynamics of infectious diseases. Ecol Lett 9, 467–484 (2006).

3. D. N. Fisman, Seasonality of infectious diseases. Annu Rev Public Health 28, 127–143 (2007).

4. D. Fisman, Seasonality of viral infections: mechanisms and unknowns. Clin Microbiol Infect 18, 946–954 (2012).

5. M. E. Martinez, The calendar of epidemics: Seasonal cycles of infectious diseases. PLoS Pathog 14, e1007327 (2018).

6. W. D. Smith, Hippocrates (Harvard University Press, 1994), vol. 7.

7. W. Cullen, The works of William Cullen: containing his physiology, nosology, and first lines of the practice of physic; with numerous extracts from his manuscript papers, and from his treatise of the materia medica (Blackwood, 1827), vol. 2.

8. A. C. Lowen, S. Mubareka, J. Steel, P. Palese, Influenza virus transmission is dependent on relative humidity and temperature. PLoS Pathog 3, 1470–1476 (2007).

9. A. C. Lowen, J. Steel, S. Mubareka, P. Palese, High temperature (30 degrees C) blocks aerosol but not contact transmission of influenza virus. J Virol 82, 5650–5652 (2008).

10. J. Shaman, M. Kohn, Absolute humidity modulates influenza survival, transmission, and seasonality. Proc Natl Acad Sci U S A 106, 3243–3248 (2009).

11. J. H. Hemmes, K. C. Winkler, S. M. Kool, Virus survival as a seasonal factor in influenza and poliomylitis. Antonie Van Leeuwenhoek 28, 221–233 (1962).

12. K. J. Reagan, M. L. McGeady, R. L. Crowell, Persistence of human rhinovirus infectivity under diverse environmental conditions. Appl Environ Microbiol 41, 618–620 (1981).

13. B. Bean et al., Survival of influenza viruses on environmental surfaces. J Infect Dis 146, 47–51 (1982).

14. Y. G. Karim, M. K. Ijaz, S. A. Sattar, C. M. Johnson-Lussenburg, Effect of relative humidity on the airborne survival of rhinovirus-14. Canadian journal of microbiology 31, 1058–1061 (1985).

15. C. M. Walker, G. Ko, Effect of ultraviolet germicidal irradiation on viral aerosols. Environ Sci Technol 41, 5460–5465 (2007).

16. J. W. Tang, The effect of environmental parameters on the survival of airborne infectious agents. J R Soc Interface 6 Suppl 6, S737–746 (2009).

17. L. M. Casanova, S. Jeon, W. A. Rutala, D. J. Weber, M. D. Sobsey, Effects of air temperature and relative humidity on coronavirus survival on surfaces. Appl Environ Microbiol 76, 2712–2717 (2010).

18. R. Grais, J. H. Ellis, A. Kress, G. Glass, Modeling the spread of annual influenza epidemics in the US: The potential role of air travel. Health care management science 7, 127–134 (2004).

19. A. Mangili, M. A. Gendreau, Transmission of infectious diseases during commercial air travel. The Lancet 365, 989–996 (2005).

20. J. S. Brownstein, C. J. Wolfe, K. D. Mandl, Empirical evidence for the effect of airline travel on inter-regional influenza spread in the United States. PLoS Med 3, e401 (2006).

21. A. T. Pavia, Germs on a plane: aircraft, international travel, and the global spread of disease. The Journal of infectious diseases 195, 621 (2007).

22. E. F. Foxman et al., Temperature-dependent innate defense against the common cold virus limits viral replication at warm temperature in mouse airway cells. Proc Natl Acad Sci U S A 112, 827–832 (2015).

23. E. Mourtzoukou, M. E. Falagas, Exposure to cold and respiratory tract infections. The International Journal of Tuberculosis and Lung Disease 11, 938–943 (2007).

24. M. E. Falagas et al., Effect of meteorological variables on the incidence of respiratory tract infections. Respiratory medicine 102, 733–737 (2008).

25. T. M. Mäkinen et al., Cold temperature and low humidity are associated with increased occurrence of respiratory tract infections. Respiratory medicine 103, 456–462 (2009).

26. J. Shaman, V. E. Pitzer, C. Viboud, B. T. Grenfell, M. Lipsitch, Absolute humidity and the seasonal onset of influenza in the continental United States. PLoS Biol 8, e1000316 (2010).

27. J. Tamerius et al., Global influenza seasonality: reconciling patterns across temperate and tropical regions. Environ Health Perspect 119, 439–445 (2011).

28. I. Chattopadhyay, E. Kiciman, J. W. Elliott, J. L. Shaman, A. Rzhetsky, Conjunction of factors triggering waves of seasonal influenza. Elife 7 (2018).

29. H. F. Dowling, G. G. Jackson, I. G. Spiesman, T. Inouye, Transmission of the common cold to volunteers under controlled conditions. III. The effect of chilling of the subjects upon susceptibility. American journal of hygiene 68, 59–65 (1958).

30. R. G. Douglas Jr, K. M. Lindgren, R. B. Couch, Exposure to cold environment and rhinovirus common cold: failure to demonstrate effect. New England Journal of Medicine 279, 742–747 (1968).

31. R. Eccles, J. Wilkinson, Exposure to cold and acute upper respiratory tract infection. Rhinology 53, 99–106 (2015).

32. S. F. Dowell, C. G. Whitney, C. Wright, C. E. Rose Jr, A. Schuchat, Seasonal patterns of invasive pneumococcal disease. Emerging infectious diseases 9, 574 (2003).

33. W. J. Alonso et al., Seasonality of Influenza in Brazil: A Traveling Wave from the Amazon to the Subtropics. American Journal of Epidemiology 165, 1434–1442 (2007).

34. E. L. Murray et al., Rainfall, household crowding, and acute respiratory infections in the tropics. Epidemiol Infect 140, 78–86 (2012).

35. K. Bloom-Feshbach et al., Latitudinal variations in seasonal activity of influenza and respiratory syncytial virus (RSV): a global comparative review. PLoS One 8, e54445 (2013).

36. J. D. Tamerius et al., Environmental predictors of seasonal influenza epidemics across temperate and tropical climates. PLoS Pathog 9, e1003194 (2013).

37. H. M. Lam et al., Nonannual seasonality of influenza-like illness in a tropical urban setting. Influenza Other Respir Viruses 12, 742–754 (2018).

38. V. G. Bruce, F. Weight, C. S. Pittendrigh, Resetting the sporulation rhythm in Pilobolus with short light flashes of high intensity. Science 131, 728–730 (1960).

39. M. C. Lobban, The entrainment of circadian rhythms in man. Cold Spring Harb Symp Quant Biol 25, 325–332 (1960).

40. M. Ishiura et al., Expression of a gene cluster kaiABC as a circadian feedback process in cyanobacteria. Science 281, 1519–1523 (1998).

41. G. W. Bartelmez, Menstruation. Journal of the American Medical Association 116, 702–704 (1941).

42. M. Fingerman, Persistent daily and tidal rhythms of color change in Callinectes sapidus. The Biological Bulletin 109, 255–264 (1955).

43. J. D. Palmer, Tidal rhythms: the clock control of the rhythmic physiology of marine organisms. Biological Reviews 48, 377–418 (1973).

44. R. Lee, Latitude and photoperiodism. Archiv für Meteorologie, Geophysik und Bioklimatologie, Serie B 18, 325–332 (1970).

45. J. L. Linzell, Innate seasonal oscillations in the rate of milk secretion in goats. J Physiol 230, 225–233 (1973).

46. D. E. Davis, Hibernation and circannual rhythms of food consumption in marmots and ground squirrels. Q Rev Biol 51, 477–514 (1976).

47. N. Mrosovsky, Circannual cycles in golden-mantled ground squirrels: phase shift produced by low temperatures. Journal of comparative physiology 136, 349–353 (1980).

48. N. Mrosovsky, Circannual cycles in golden-mantled ground squirrels: fall and spring cold pulses. Journal of Comparative Physiology A 167, 683–689 (1990).

49. G. A. Lincoln, H. Andersson, D. Hazlerigg, Clock genes and the long-term regulation of prolactin secretion: evidence for a photoperiod/circannual timer in the pars tuberalis. J Neuroendocrinol 15, 390–397 (2003).

50. G. A. Lincoln, I. J. Clarke, R. A. Hut, D. G. Hazlerigg, Characterizing a mammalian circannual pacemaker. Science 314, 1941–1944 (2006).

51. M. Wikelski et al., Avian circannual clocks: adaptive significance and possible involvement of energy turnover in their proximate control. Philos Trans R Soc Lond B Biol Sci 363, 411–423 (2008).

52. G. Lincoln, A brief history of circannual time. J Neuroendocrinol 31, e12694 (2019).

53. S. F. Dowell, Seasonal variation in host susceptibility and cycles of certain infectious diseases. Emerg Infect Dis 7, 369–374 (2001).

54. S. Dowell, Seasonality–still confusing. Epidemiology & Infection 140, 87–90 (2012).

55. L. Hansen, The Truven health MarketScan databases for life sciences researchers. Truven Health Ananlytics IBM Watson Health (2017).

56. M. Sengupta et al., The national solar radiation data base (NSRDB). Renewable and Sustainable Energy Reviews 89, 51–60 (2018).

57. G. B. Ermentrout, J. Rinzel, Beyond a pacemaker’s entrainment limit: phase walk-through. American Journal of Physiology-Regulatory, Integrative and Comparative Physiology 246, R102–R106 (1984).

58. S. M. Cook, R. I. Glass, C. W. LeBaron, M. S. Ho, Global seasonality of rotavirus infections. Bull World Health Organ 68, 171–177 (1990).

59. L. P. Shek, B. W. Lee, Epidemiology and seasonality of respiratory tract virus infections in the tropics. Paediatr Respir Rev 4, 105–111 (2003).

60. M. Pons-Salort et al., The seasonality of nonpolio enteroviruses in the United States: Patterns and drivers. Proc Natl Acad Sci U S A 115, 3078–3083 (2018).

61. W. P. London, J. A. Yorke, Recurrent outbreaks of measles, chickenpox and mumps. I. Seasonal variation in contact rates. Am J Epidemiol 98, 453–468 (1973).

62. K. Dietz, “The incidence of infectious diseases under the influence of seasonal fluctuations” in Mathematical models in medicine. (Springer, 1976), pp. 1–15.

63. M. Pascual, X. Rodó, S. P. Ellner, R. Colwell, M. J. Bouma, Cholera Dynamics and El Niño-Southern Oscillation. Science 289, 1766–1769 (2000).

64. J. Dushoff, J. B. Plotkin, S. A. Levin, D. J. Earn, Dynamical resonance can account for seasonality of influenza epidemics. Proceedings of the National Academy of Sciences 101, 16915–16916 (2004).

65. N. C. Grassly, C. Fraser, G. P. Garnett, Host immunity and synchronized epidemics of syphilis across the United States. Nature 433, 417–421 (2005).

66. M. Greer, R. Saha, A. Gogliettino, C. Yu, K. Zollo-Venecek, Emergence of oscillations in a simple epidemic model with demographic data. R Soc Open Sci 7, 191187 (2020).

67. M. Vyska, C. Gilligan, Complex Dynamical Behaviour in an Epidemic Model with Control. Bull Math Biol 78, 2212–2227 (2016).

68. T. G. Paglieroni, P. V. Holland, Circannual variation in lymphocyte subsets, revisited. Transfusion 34, 512–516 (1994).

69. R. J. Nelson, G. E. Demas, Seasonal changes in immune function. Q Rev Biol 71, 511–548 (1996).

70. A. Tendler et al., Hormone seasonality in medical records suggests circannual endocrine circuits. Proceedings of the National Academy of Sciences 118, e2003926118 (2021).

71. Anonymous, U.S. Census Bureau 2010 Census Available at:https://www.census.gov/2010census/data/.

72. P. Virtanen et al., SciPy 1.0: fundamental algorithms for scientific computing in Python. Nature methods 17, 261–272 (2020).

73. S. Seabold, J. Perktold (2010) Statsmodels: Econometric and statistical modeling with python. in Proceedings of the 9th Python in Science Conference (Austin, TX), p 61.

