## Supplemental Text for "Seasonal Disease in the United States Has the Hallmarks of an Entrained Circannual Clock"

**This PDF file includes:**

Supplementary text
Figure S1
Tables S1 to S2
Legend for Dataset S1

**Other supplementary materials for this manuscript include the following:**

Dataset S1

**Supplementary Information Text**

**Details of the driven oscillator model.**

When forcing is weak, one can approximate a generic driven oscillator with linearized dynamics using a second-order Taylor series expansion. The result is a familiar equation from classical mechanics which describes the behavior of damped harmonic oscillators, a physical analogy of which being a swing-set or pendulum. The second-order linear differential equation is as follows:

$$\frac{d^2x}{\mathrm{d}t^2} = -{\omega_0}^2 x - \zeta\omega_0 \frac{dx}{dt} + A\sin(\Omega t)$$

For natural frequency $\omega_0$, dampening ratio $\zeta$, driving amplitude $A$, and driving frequency $\Omega$. The response of the system has the following analytical solution:

$$x(t) = X_0 \sin(\Omega t + \theta)$$

Where the phase of the system $\theta$ is described by:

$$\theta = \arctan\left(\frac{\zeta\omega_0\Omega}{{\omega_0}^2 - \Omega^2}\right)$$

And the amplitude $X_0$ has the form:

$$X_0 = \frac{A}{\sqrt{({\omega_0}^2 - \Omega^2)^2 + \zeta^2{\omega_0}^2\Omega^2}}$$

Several insights can be gained from these expressions. First, this type of system will always oscillate in sync with the environment, no matter how weak the driving amplitude $A$, or what frequency $\Omega$. Second, the phase of the system $\theta$ is not dependent on driving amplitude $A$. In contrast, the amplitude of such systems scales proportionally with changes in driving strength. For the purpose of modeling disease time-series, we replaced the input expression $A\sin(\Omega t)$ with an arbitrary linear combination of weather time-series:

$$\mathrm{input}(t) = \sum_{i=1}^{n} a_i \mathrm{var}_i(t)$$

Where $\mathrm{var}_i(t)$ denote weather time-series obtained individually for each county and scalar coefficients $a_i$ represent global weightings of each possible driving force. Here, the inputs $i = (1, \ldots, n)$ are comprised of daily temperature, solar irradiance, and specific humidity. Finally, the driven response $x(t)$ was allowed to relate to the final output, seasonal infection incidence rate, through the following linear function with temporal lag $\tau$:

$$\mathrm{output}(t) = \alpha x(t-\tau) + \beta$$

For simplicity, the frequency of the environment $\Omega$ was assumed to equal $2\pi/365$. Thus, nonlinear regression could be used to find parameters $\omega_0, \zeta, \alpha, \beta, \tau, a_i$ that most accurately relate the input signal $\mathrm{input}(t)$ to time-series of seasonal disease in each county.

**Details of the phase oscillator model.**

The simplest model of a weakly driven limit cycle is parameterized only by its phase $\theta$, where the driving force is effectively always tangential to the orbit. The coupling between the limit cycle oscillator and its driving force can be any periodic function of the phase mismatch $\phi - \theta$. From Ermentrout and Rinzel (58) we have the simplest possibility:

$$\frac{d\theta}{dt} = \omega + A\sin(\phi - \theta)$$

$$\frac{d\phi}{dt} = \Omega$$

Where $\omega$ is the natural frequency of the limit cycle, $\phi$ denotes the phase of the driving force, $A$ is the driving amplitude, and $\Omega$ is the frequency of the driving signal. One can learn about the response of the system by considering only phase locked solutions in which the angle between the phase oscillator and the environment $\phi - \theta$ is constant:

$$0 = \frac{d}{dt}(\phi - \theta) = \Omega - \omega - A\sin(\phi - \theta)$$

By solving for $\phi - \theta$ we obtain:

$$\phi - \theta = \arcsin\left(\frac{\Omega - \omega}{A}\right)$$

This equation has several implications. First, entrainment of the system requires the argument of $\arcsin$ to be less than 1. Thus, a phase oscillator will only synchronize with its environment when the driving amplitude $A$ is sufficiently large compared to its frequency mismatch with the environment $\Omega - \omega$. The relative phase of entrained solutions varies inversely with driving amplitude like $\sim 1/A$. For the purpose of modeling disease time-series, we expanded the input expression to include a county-specific amplitude term $\gamma$, yielding: $A\gamma\sin(\phi - \theta)$. Similar to our treatment of the driven oscillator, county-specific input amplitude $\gamma$ and phase $\phi(t)$ were inferred from a linear combination of weather time-series $\mathrm{var}_i(t)$, parameterized by scalar coefficients $a_i$:

$$\mathrm{input}(t) = \sum_{i=1}^{n} a_i \mathrm{var}_i(t)$$

The entraining cue $\mathrm{input}(t)$ was defined individually for each county based on their weather time-series. For simplicity, the frequency of the environment $\Omega$ was assumed to equal $2\pi/365$. Lastly, the output of the system was taken as a simple sinusoidal function of the phase $\theta(t)$:

$$\mathrm{output}(t) = \alpha \sin(\theta(t) - \Theta) + \beta$$

Because the long-term behavior of stably entrained solutions depends only on the ratio $(\Omega - \omega)/A\gamma$, the system is degenerate with respect to the frequency mismatch and driving amplitude $A\gamma$, which cannot be known exactly. In modeling disease time-series, we fixed $\omega = 2\pi/300$ while parameters $A, \alpha, \beta, \Theta, a_i$ were fit through nonlinear regression.

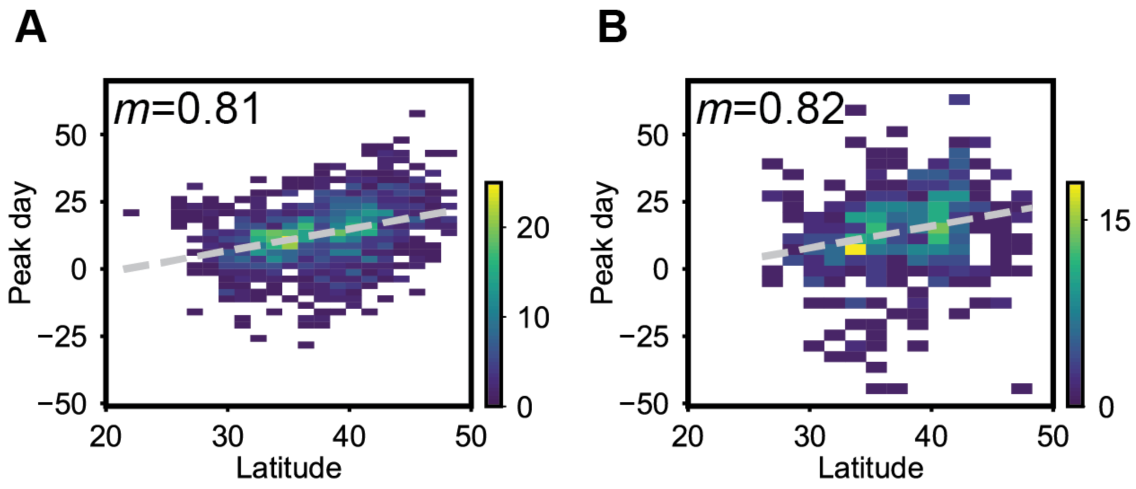


**Fig. S1. (A)** Latitude versus peak timing for an aggregate of less common diagnoses. To assess the robustness of the latitude-phase scaling, the top 10 most common diagnoses, comprising over 70% of the dataset (see Table S1) were excluded. After filtering based on noise criteria (see Materials and Methods), a total of 1,183 counties were included. The resulting linear slope ($\boldsymbol{m}$) and corresponding Pearson correlation coefficient ($\boldsymbol{R = 0.35, p < 2.55 \times 10^{-34}}$) are consistent with what is observed in the main analysis. **(B)** Latitude versus peak timing for Streptococcal Sore Throat, the most common diagnosis code associated with a singular pathogen. After filtering based on noise criteria (see Materials and Methods), a total of 476 counties were included in the analysis. The resulting linear slope ($\boldsymbol{m}$) and corresponding Pearson correlation coefficient ($\boldsymbol{R = 0.23, p < 3.53 \times 10^{-07}}$) are consistent with the main analysis.

**Table S1.** Diagnoses included in this study.

| Identifier[a] | Diagnosis description | Percent of total |
|---|---|---|
| 4659 | Acute upper respiratory infections of unspecified site | 13.02 |
| 462 | Acute pharyngitis | 12.14 |
| 4619 | Acute sinusitis, unspecified | 10.16 |
| 4660 | Acute bronchitis | 8.01 |
| 7862 | Cough | 7.79 |
| 3829 | Unspecified otitis media | 5.89 |
| 486 | Pneumonia, organism unspecified | 4.60 |
| 4739 | Unspecified sinusitis (chronic) | 3.45 |
| 07999 | Unspecified viral infection | 3.11 |
| 38200 | Acute suppurative otitis media without spontaneous rupture of eardrum | 3.05 |
| 0340 | Streptococcal sore throat | 2.94 |
| 490 | Bronchitis, not specified as acute or chronic | 2.32 |
| 4610 | Acute maxillary sinusitis | 1.86 |
| 463 | Acute tonsillitis | 1.57 |
| 38181 | Dysfunction of Eustachian tube | 1.43 |
| 38870 | Otalgia, unspecified | 1.25 |
| 78703 | Vomiting alone | 1.20 |
| 460 | Acute nasopharyngitis [common cold] | 1.10 |
| 49121 | Obstructive chronic bronchitis with (acute) exacerbation | 1.06 |
| 38101 | Acute serous otitis media | 0.87 |
| 78607 | Wheezing | 0.81 |
| 4730 | Chronic maxillary sinusitis | 0.80 |
| 38110 | Chronic serous otitis media, simple or unspecified | 0.79 |
| 46619 | Acute bronchiolitis due to other infectious organisms | 0.69 |
| 4658 | Acute upper respiratory infections of other multiple sites | 0.68 |
| 0088 | Intestinal infection due to other organism, not elsewhere classified | 0.63 |
| 4618 | Other acute sinusitis | 0.56 |
| 38100 | Acute nonsuppurative otitis media, unspecified | 0.55 |
| 3814 | Nonsuppurative otitis media, not specified as acute or chronic | 0.50 |
| 4611 | Acute frontal sinusitis | 0.48 |
| 4738 | Other chronic sinusitis | 0.41 |
| 7841 | Throat pain | 0.36 |
| 4829 | Bacterial pneumonia, unspecified | 0.35 |
| 075 | Infectious mononucleosis | 0.29 |
| 46400 | Acute laryngitis without mention of obstruction | 0.28 |
| 7068 | Other specified diseases of sebaceous glands | 0.26 |
| 5198 | Other diseases of respiratory system, not elsewhere classified | 0.25 |
| 0091 | Colitis, enteritis, and gastroenteritis of presumed infectious origin | 0.25 |
| 4650 | Acute laryngopharyngitis | 0.24 |

| Identifier[a] | Diagnosis description | Percent of total |
|---|---|---|
| 3813 | Other and unspecified chronic nonsuppurative otitis media | 0.23 |
| 5110 | Pleurisy without mention of effusion or current tuberculosis | 0.22 |
| 51884 | Acute and chronic respiratory failure | 0.19 |
| 7869 | Other symptoms involving respiratory system and chest | 0.19 |
| 481 | Pneumococcal pneumonia [Streptococcus pneumoniae pneumonia] | 0.17 |
| 37203 | Other mucopurulent conjunctivitis | 0.17 |
| 3823 | Unspecified chronic suppurative otitis media | 0.16 |
| 4919 | Unspecified chronic bronchitis | 0.15 |
| 4910 | Simple chronic bronchitis | 0.14 |
| 485 | Bronchopneumonia, organism unspecified | 0.14 |
| 38120 | Chronic mucoid otitis media, simple or unspecified | 0.14 |
| 0090 | Infectious colitis, enteritis, and gastroenteritis | 0.14 |
| 4731 | Chronic frontal sinusitis | 0.13 |
| 38871 | Otogenic pain | 0.13 |
| 5362 | Persistent vomiting | 0.12 |
| 683 | Acute lymphadenitis | 0.11 |
| 4721 | Chronic pharyngitis | 0.11 |
| 5199 | Unspecified disease of respiratory system | 0.11 |
| 3824 | Unspecified suppurative otitis media | 0.11 |
| 38201 | Acute suppurative otitis media with spontaneous rupture of eardrum | 0.11 |
| 49122 | Obstructive chronic bronchitis with acute bronchitis | 0.10 |
| 4612 | Acute ethmoidal sinusitis | 0.09 |
| 38860 | Otorrhea, unspecified | 0.09 |
| 78031 | Febrile convulsions (simple), unspecified | 0.09 |
| 4809 | Viral pneumonia, unspecified | 0.08 |
| 07799 | Unspecified diseases of conjunctiva due to viruses | 0.08 |
| 48289 | Pneumonia due to other specified bacteria | 0.08 |
| 4789 | Other and unspecified diseases of upper respiratory tract | 0.07 |
| 0341 | Scarlet fever | 0.07 |
| 0796 | Respiratory syncytial virus (RSV) | 0.06 |
| 4838 | Pneumonia due to other specified organism | 0.06 |
| 51919 | Other diseases of trachea and bronchus | 0.06 |
| 7908 | Viremia, unspecified | 0.06 |
| 07989 | Other specified viral infection | 0.06 |

**Table S2.** Best fit parameters for models of disease incidence time-series.

**Driven oscillator model parameters**

| Core oscillator (1/day) | | Output function | | | Input function | | |
|---|---|---|---|---|---|---|---|
| **ω** | **ζ** | **α** | **β** | **τ** | $a_{temperature}$ | $a_{irradiance}$ | $a_{humidity}$ |
| 0.0163 | 0.0219 | 37.47 | 1865.50 | 37.04 | $2.21 \times 10^{-4}$ | $3.97 \times 10^{-6}$ | $2.97 \times 10^{-4}$ |

**Phase oscillator model parameters**

| Core oscillator (1/day) | | Output function | | | Input function | | |
|---|---|---|---|---|---|---|---|
| **A** | **ω** (fixed) | **α** | **β** | **Θ** | $a_{temperature}$ | $a_{irradiance}$ | $a_{humidity}$ |
| 2.56 | 0.021 | 798.68 | 1950.29 | 5.58 | $1.00 \times 10^{-10}$ | $2.94 \times 10^{-3}$ | $5.23 \times 10^{-10}$ |

**Dataset S1 (separate file).** Summary statistics of seasonal disease by county.
